# IMPROVING CARDIOVASCULAR DISEASE RISK PREDICTION WITH MACHINE LEARNING USING MENTAL HEALTH DATA: A PROSPECTIVE UK BIOBANK STUDY

**DOI:** 10.1101/2022.10.23.22281428

**Authors:** Mohsen Dorraki, Zhibin Liao, Derek Abbott, Peter J. Psaltis, Emma Baker, Niranjan Bidargaddi, Hannah R. Wardill, Anton van den Hengel, Jagat Narula, Johan W. Verjans

## Abstract

**Background:** Robust and accurate prediction of cardiovascular disease (CVD) risk facilitates early intervention to benefit patients. It is well-known that mental disorders and CVD are interrelated. Nevertheless, psychological factors are not considered in existing models, which use either a limited number of clinical and lifestyle factors, or have been developed on restricted population subsets.

**Objectives:** To assess whether inclusion of psychological data could improve CVD risk prediction in a new machine learning (ML) approach.

**Methods:** Using a comprehensive, long-term UK Biobank dataset (n=375,145), we examined the correlation between CVD diagnoses and traditional and psychological risk factors. An ensemble ML model containing five constituent algorithms [decision tree, random forest, XGBoost, support vector machine (SVM), and deep neural network (DNN)] was tested for its ability to predict CVD risk based on two training datasets: one using traditional CVD risk factors alone, or a combination of traditional and psychological risk factors.

**Results:** Our ensemble ML model could predict CVD with 71.31% accuracy using traditional CVD risk factors alone. However, by adding psychological factors to the training data, accuracy dramatically increased to 85.13%. The accuracy and robustness of our ensemble ML model outperformed all five constituent learning algorithms. Re-testing the model on a control dataset to predict bone diseases returned random results, confirming specificity of the training data for prediction of CVD.

**Conclusions:** Incorporating mental health assessment data within an ensemble ML model results in a significantly improved, highly accurate, state-of-the-art CVD risk prediction.

**AUTHOR APPROVAL:** All authors have seen and approved the manuscript.

**COMPETING INTERESTS:** The authors declare no competing interests.

**DATA AVAILABILITY STATEMENT:** All data needed to evaluate the conclusions in the paper are present in the paper or in the supplementary materials. In addition, we used UK Biobank in this study: www.ukbiobank.ac.uk.

**FUNDING:** No funding.

## INTRODUCTION

Coronary heart disease and cerebrovascular disease are the world’s leading cause of death (1). Early intervention is highly effective for people at risk of developing the disease, and a widely used approach is to screen at-risk populations (2). Several Cardiovascular disease (CVD) risk prediction models have been developed during the past four decades. The Reynolds Risk Score in women, used in the US, is based on data from the Women’s Health Study (3). Another multi-marker risk model assesses the contribution of ten genetic markers, including B-type natriuretic peptide and C-reactive protein (4). SCORE, introduced by the European Society of Cardiology, was developed retrospectively from data originating from 12 European cohorts undergoing baseline examination (5), and QRISK is recommended in Europe (6,7).

Most risk prediction models follow a statistical regression approach that relates CVD risk with an individual’s characteristics (8), but the number of factors used for prediction is often limited. New approaches, covering a more comprehensive range of markers that do not rely solely on traditional clinical and lifestyle factors, could improve precision in identifying CVD risk.

In addition to traditional, recognized risk factors for CVD, such as smoking, high cholesterol levels, hypertension, and obesity, there is also evidence of a strong association between psychological factors and CVD (9). The association is bidirectional, in that psychological factors may be common in certain CVDs and portend worse outcomes, regardless of whether the psychological disorder or CVD occurs first (10).

The important role of psychological stress has been investigated in myocardial ischemia (11), coronary artery disease (12,13), acute and reversible cardiomyopathy (14), and ventricular arrhythmias (15). Prospective evidence implies that depression plays a major role CVD development (16,17). The association of depression is also reported with coronary artery disease (18,19), heart rate reactivity (20), myocardial infarction (21), and respiratory sinus arrhythmia fluctuation (22). A considerable number of studies have shown that anxiety is an independent risk factor for cardiac mortality and coronary heart disease (23–25).

Although the association between psychological factors and CVD is well documented, no method has previously been devised for exploiting the former in predicting the latter. Until now, the development of such a model has not been possible due to the lack of an appropriate dataset and an automated assessment framework. This study aims to develop an ensemble machine learning (ML) model to predict CVD risk, initially using only traditional risk factors factors [such as body mass index (BMI), hypertension (HT), systolic blood pressure (SBP), diastolic blood pressure (DBP), diabetes, hyperlipidemia, smoking status, age, and gender], then with the addition of psychological data [such as depression, anxiety, and stress], to train and assess the accuracy of our ML model. We develop an ensemble machine learning (ML) model to predict CVD risk, initially using only traditional risk factors, then with the addition of psychological data, to train and assess the accuracy of our ML model.

## METHODS

### Study design

The UK Biobank is a large-scale biomedical database and research resource containing in-depth genetic and health information. It comprises a large, long-term (since 2006) dataset from the United Kingdom, recruiting >500,000 participants from the general population—design and participant population are described in (26). Data is collected through a combination of questionnaires, interviews, and blood, urine, and saliva samples and describes these physical measurements, together with lifestyle, medical history, and a series of health risk factors.

We considered two categories as training data for our ML model. The first consisted of traditional CVD risk factors, such as gender, age, smoking status, BMI, HT, diabetes, SBP, DBP, and hyperlipidemia. The second category contained data from the UK Biobank touchscreen questionnaire on psychological factors and mental health including diagnosed disorders (bipolar disorder, depression, anxiety, etc.), and symptoms (neuroticism, mood swings, miserableness, irritability, sensitivity, hurt feelings, nervous feelings, loneliness, isolation, guilty feelings, severity of mania, etc.; full details and ICD-10 codes in Supplemental Table 1).

CVDs considered in this study included atrial fibrillation/flutter, ventricular arrhythmias, angina pectoris, acute and subsequent myocardial infarctions and subsequent complications, chronic and acute ischemic heart diseases, and hypertensive heart disease (full details in Supplemental Table 2).

After preprocessing to remove entries with incomplete data, the data from n=375,145 subjects were analyzed using a Pearson correlation between input features {*X*_1_, *X*_2_, *X*_3_, …, *X*_*n*_} and output label *y*_*i*_, where *i* = 1, 2, 3, …, *n* was mathematically defined as (27):

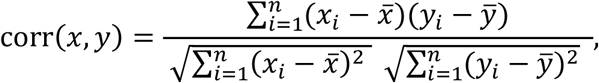

where *x* and *y* denote the input variable and the dependent variable or target feature in the dataset, respectively.

The median diagnosis age of included patients was 58.0 years (interquartile range: 37–73 years). We separated the data into training and test sets with 90% and 10% of subjects.

### Modeling and predictive analyses

We developed an ensemble model containing five ML methods: decision tree, random forest, XGBoost, support vector machine (SVM), and deep neural networks (DNN). Briefly, a decision tree algorithm (28) employs a tree-like model of decisions and their possible consequences, including chance event outcomes, resource costs, and utility. Random forest (29) is a learning method operating by constructing a multitude of decision trees at training time and XGBoost (30) is a scalable end-to-end tree boosting system. An SVM (31) is a supervised ML approach that maps training examples to points in space to maximize the width of the gap between the two clusters of points. A neural network recognizes hidden patterns in a dataset by mimicking the human brain, and DNNs (32) possess multiple layers to progressively extract higher-level patterns from raw input.

Our proposed ensemble-learning model collected results from each individual learning approach and produced a final output prediction by adopting a majority voting strategy; each model makes a prediction (votes) for each test instance and the final result is the one that receives more than half the votes. The process of UK Biobank data collection and our model overview are shown in the Figure 1.

**Figure 1:**
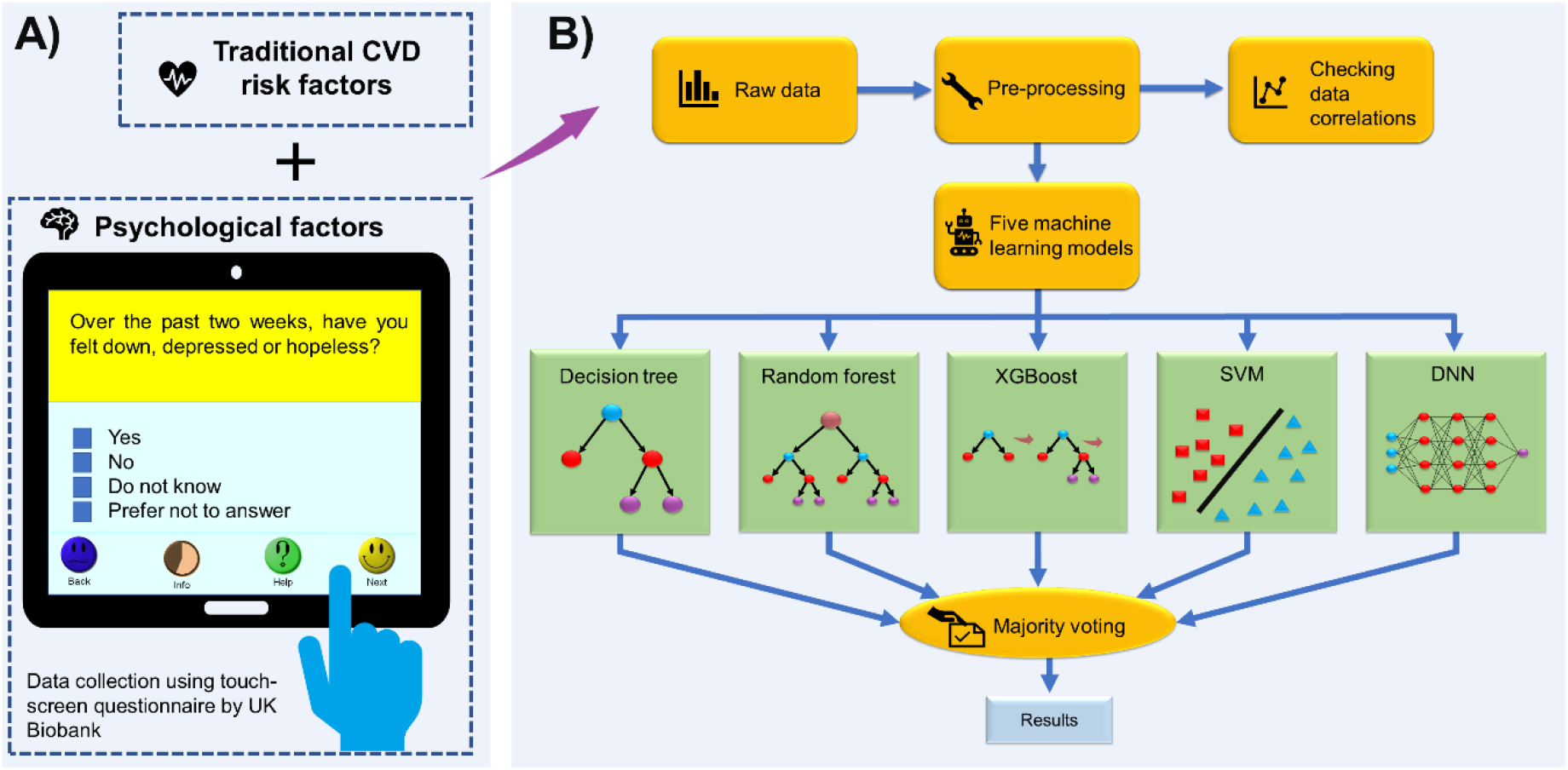
Model overview. A) Data collection by the UK Biobank. B) Our ensemble machine learning approach, including decision tree, random forest, XGBoost, SVM, and DNN models, was developed to predict CVD risk from psychological factors.

## RESULTS

### Risk factor correlation

Our correlation analysis (Figure 2A) shows that almost all psychological factors are highly correlated, with the exception of risk taking (code:2040). There is generally a negative correlation between age and psychological disorders, which may imply that psychological illnesses are not a natural part of ageing, and that psychological disorders predominantly affect younger adults.

**Figure 2:**
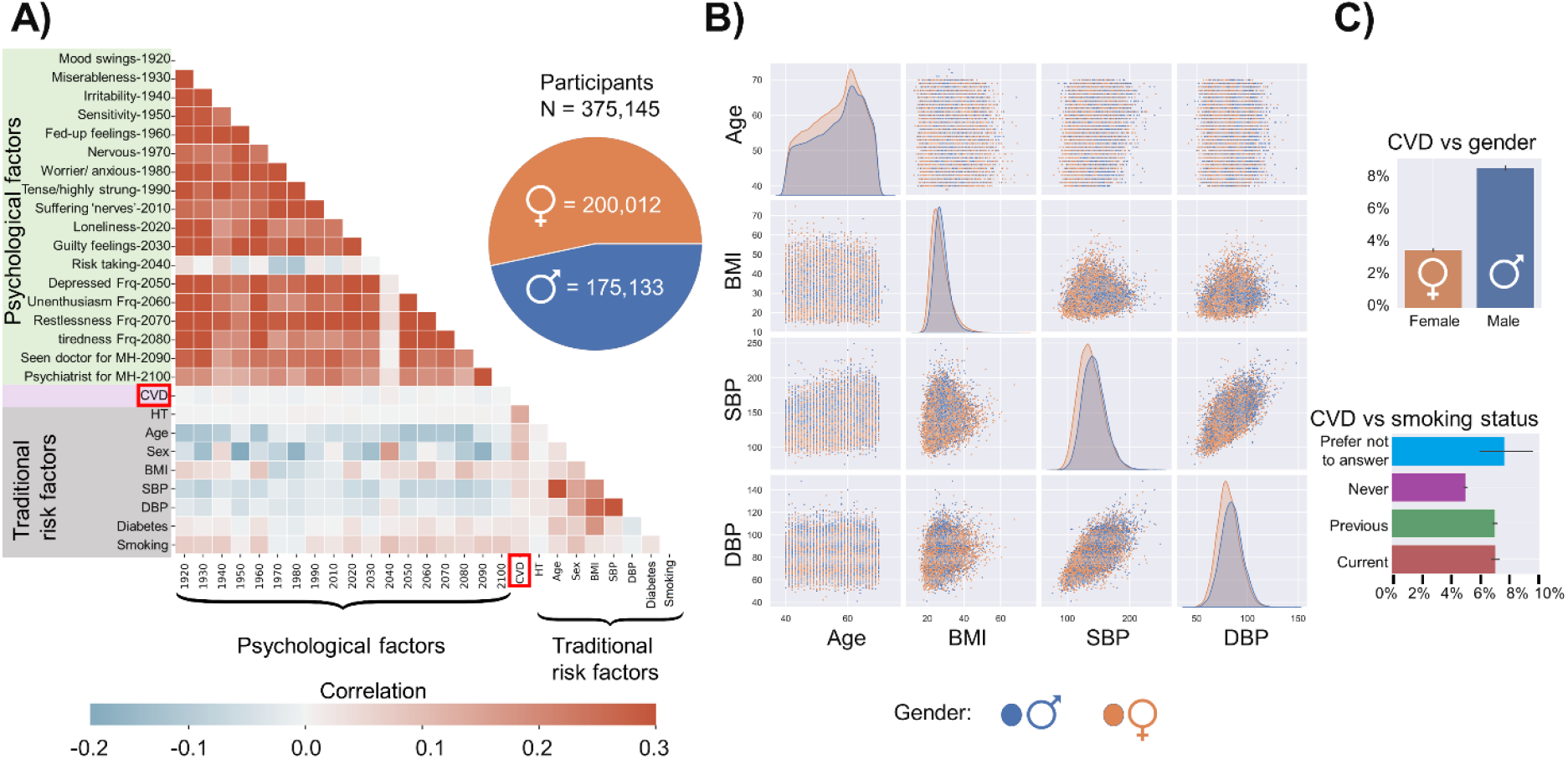
CVD risks for participants from the UK Biobank. A) The correlation between CVD and a selection of psychological factors and traditional CVD risk factors. Descriptions for psychological factor codes are provided in Supplemental Table 1. B) The association of four traditional CVD risk factors by gender: age, BMI, SBP, and DBP. C) The prevalence of CVD by gender and smoking status.

Positive associations were observed between CVD and the traditional risk factors, especially for HT (Figure 2A). Figure 2B depicts the distribution of key physical factors (age, BMI, SBP, DBP) by gender. The incidence of CVD in females is lower than in males, and CVD incidence increases for previous and current smokers (Figure 2C).

### Prediction of CVD

We explored the ability of our model to predict CVD from two different groups of training data, namely traditional CVD risk factors only, and the combination of traditional and psychological factors. We separately trained the model on each dataset, then evaluated its accuracy with unseen test data.

Trained using traditional CVD risk factors only (gender, age, BMI, HT, hyperlipidemia, SBP, DBP, diabetes, and smoking status), our ensemble model was able to predict CVD with an accuracy of 71.31% (precision 74.91%; recall 70.48%), which exceeded the performance of each individual ML model, particularly in precision (Figure 3A; metrics defined in Supplemental Table 3).

**Figure 3.**
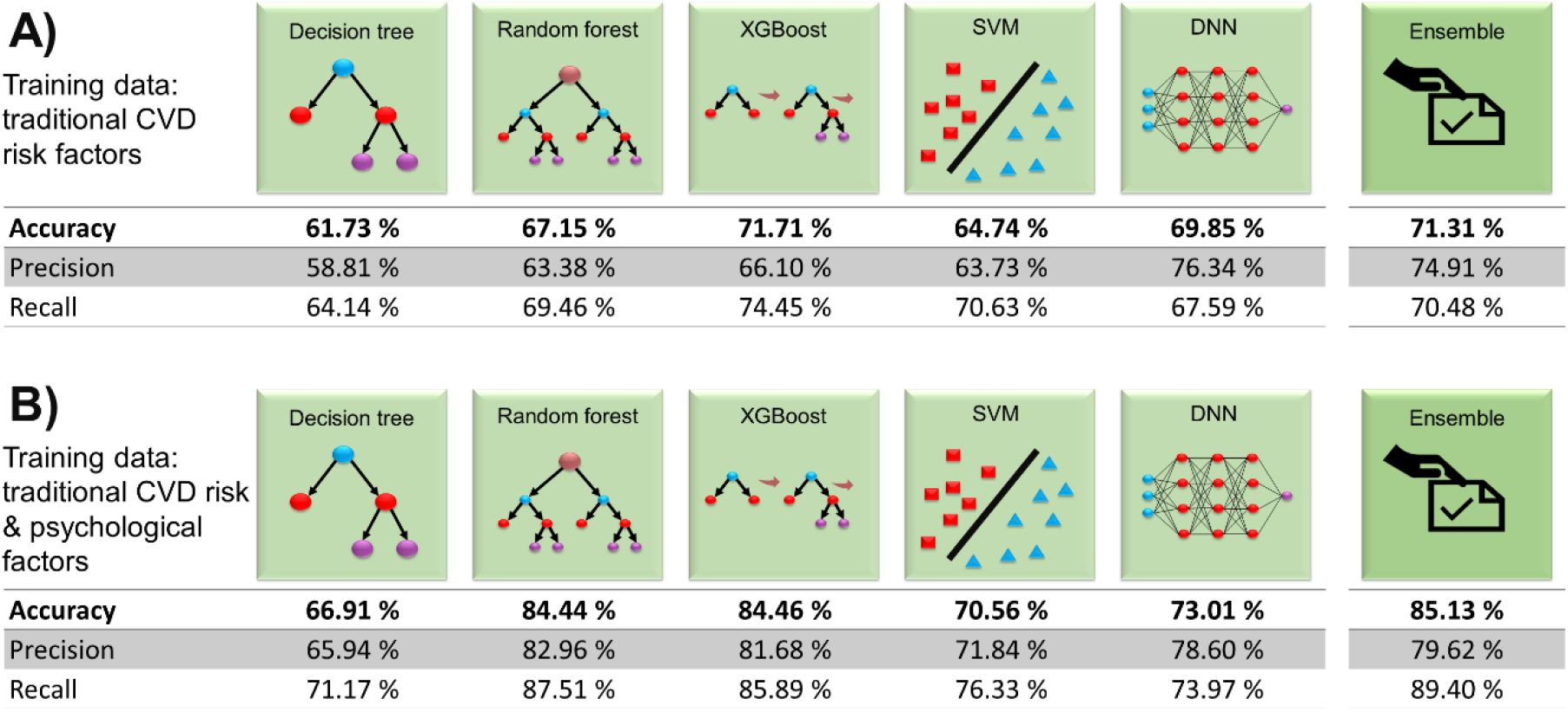
Results from different ML models for two training data categories. Accuracy, precision, and recall results from our ensemble model and five individual learning approaches are shown for training data comprising A) traditional CVD risk factors only; and B) the combination of traditional CVD risk and psychological factors. All metrics are described in Supplemental Table 3.

A second, independent training using combined traditional and psychological risk factors significantly increased CVD prediction accuracy of the ensemble model to 85.13% (precision 79.62%; and recall 89.40%; Figure 3B). The accuracy and recall of our ensemble model outperformed any of the five constituent learning algorithms; intriguingly, the precision of the individual random forest and DNN algorithms were higher than for the ensemble model, suggesting the appropriate data classification of the ensemble model.

Thus, adding amassed psychological data to train our ensemble model increased the baseline 71.31% accuracy of CVD risk prediction by over 10%, to be 85.13% accurate. All other performance metrics also increased significantly after adding psychological factors to the training data, indicating their value in improving identification of patients at-risk of CVD.

### Bone disease prediction cross-check

To examine whether the predicted results were not generated spuriously, we performed a control experiment using the UK Biobank bone disease dataset. Using our model trained on traditional and psychological CVD risk factors, we tested its accuracy in predicting bone diseases such as tuberculosis of bones and joints, disorders of bone density and structure, and osteopathies (see Supplemental Table 4). The ensemble model returned performance results similar to those of the five individual approaches; none were effective at predicting bone disorders using CVD risk factors (Supplemental Figure 1).

## DISCUSSION

In this prospective study, we developed an ensemble ML approach that can accurately predict CVD risk from traditional risk factors alone. The accuracy of CVD prediction substantially improved to more than >85% by including mental health assessment data in the model, which addressed participant symptoms and disorders such as depression, anxiety, and stress.

Prediction of CVD risk is a critical domain within CVD prevention. Several CVD risk prediction systems, such as SCORE (5), work with traditional risk factors, e.g., age, sex, smoking, SBP, and total cholesterol. While these factors correlate well with CVD, their number is limited. Further, some prediction systems have been developed on narrow datasets, e.g., subjects with age of 40–65 years for SCORE and 40–69 years for SCORE2, limiting their validity to subjects outside that range.

Because of the relatively limited performance, and variation on individual level, new approaches are needed to guide primary prevention with features that do not rely solely on a limited number of clinical and lifestyle factors, and that can be applied to a broader range of individuals. Psychological studies have shown that mental health factors can be used for CVD risk prediction, which was demonstrated via correlation analysis and statistical risk prediction (33). In this study we have used for the first time an ML approach that exploits these relationships to improve CVD risk prediction to impact management and early intervention.

ML approaches have the capability to interpret hidden patterns and structures within psychological and CVD data, that cannot be detected by humans or traditional statistical approaches. These approaches have the power to analyze very large quantities of data to generate data-driven recommendations, rather than hypothesis driven answers. These intelligent methods do not discount the value of conventional diagnosis assessments to evaluate CVD risks; rather, they provide opportunities to predict or make diagnoses faster, more convenient, and more accurate by analyzing larger volumes of data than would be possible by humans.

Over the past few years, ML researchers have studied ensemble ML schemes that employ multiple approaches to provide more accurate performance than could be returned from any single constituent learning algorithm. In this study, we have developed an ensemble ML model to analyze UK Biobank data that improved the decision robustness and accuracy over any individual approach.

For the first time, we show that adding mental health factors to traditional CVD risk factors can improve the accuracy of ML prediction models. Our results may open new avenues for medical researchers and clinicians to predict CVD risk more precisely and apply early intervention strategies for at-risk individuals using a single time point mental health assessment. The principles demonstrated here can be readily applied to more complex ML models with more input data. Comprehensive biobank datasets are necessary for training and validation, and these are becoming more available in healthcare settings through several sources such as hospitals. Our ensemble ML approach can be used to automate analysis and management of these big datasets to derive meaningful information to benefit clinical outcomes.

### Study limitations

We have focused on the prediction of CVD using traditional and psychological factors regardless of CVD diagnosis timing. Although the date of first CVD diagnosis and the date of completing mental health questionnaires are available for individual patients, it is difficult to conclude with certainty whether the reported psychological disorders preceded or followed the development of CVD, especially as the mental health questionnaires only began in August 2016 (Supplemental Figure 2). Addressing this question is challenging as many patients reported with psychological disorders have unknowingly already developed CVD or vice versa. Investigations typically assume that the personality structure occurred before the appearance of heart disease (34), however, a longitudinal study of the personality of individuals prior to CVD would be required to address this question, opening future opportunities to investigate causal relationships.

## CONCLUSIONS

We have developed a new CVD risk prediction ensemble ML model that improves CVD prediction outcomes relative to all five constituent algorithms when trained on traditional CVD risk factors. The accuracy remarkably increased to more than 85% when the model was trained using mental health data in addition to traditional CVD risk factor data. The findings imply that psychological assessment can become a reliable, easy and affordable contribution to improving CVD risk prediction and management using ML approaches.

## Data Availability

All data needed to evaluate the conclusions in the paper are present in the paper or in the supplementary materials. In addition, we used UK Biobank in this study: www.ukbiobank.ac.uk.

