## Supplementary material for "IMPROVING CARDIOVASCULAR DISEASE RISK PREDICTION WITH MACHINE LEARNING USING MENTAL HEALTH DATA: A PROSPECTIVE UK BIOBANK STUDY": Psy&CVD.v12.Supplemental Material.pdf

**Supplementary Material** | **Page**


| Supplementary Material | Page |
|---|---|
| **Supplemental Table 1.** List of psychological factors and their UK Biobank codes used in this study. | 2 |
| **Supplemental Table 2.** CVDs and corresponding ICD-10 codes used in this study. | 3 |
| **Supplemental Table 3.** Definition of ML performance metrics. | 4 |
| **Supplemental Table 4.** Bone diseases and corresponding ICD-10 codes used in this study. | 5 |
| **Supplemental Figure 1.** ML model predictions of bone disease from CVD training data. | 7 |
| **Supplemental Figure 2.** Date of first CVD in-patient diagnosis versus date of completing UK Biobank mental health questionnaire (started in August 2016). | 8 |

**Supplemental Table 1.** List of psychological factors and their UK Biobank codes used in this study.

| Field ID | Description |
|---|---|
| 1920 | Mood swings |
| 1930 | Miserableness |
| 1940 | Irritability |
| 1950 | Sensitivity/ hurt feelings |
| 1960 | Fed-up feelings |
| 1970 | Nervous feelings |
| 1980 | Worrier / anxious feelings |
| 1990 | Tense / 'highly strung' |
| 2000 | Worry too long after embarrassment |
| 2010 | Suffer from 'nerves' |
| 2020 | Loneliness, isolation |
| 2030 | Guilty feelings |
| 2040 | Risk taking |
| 2050 | Frequency (Frq) of depressed mood in last 2 weeks |
| 2060 | Frequency of unenthusiasm / disinterest in last 2 weeks |
| 2070 | Frequency of tenseness / restlessness in last 2 weeks |
| 2080 | Frequency of tiredness / lethargy in last 2 weeks |
| 2090 | Seen doctor (GP) for nerves, anxiety, tension, or depression (mental health) |
| 2100 | Seen a psychiatrist for nerves, anxiety, tension, or depression |
| 20126 | Bipolar and major depression status |
| 20127 | Neuroticism score |
| 20124 | Probable recurrent major depression (moderate) |
| 20125 | Probable recurrent major depression (severe) |
| 20123 | Single episode of probable major depression |
| 4598 | Ever depressed for a whole week |
| 4609 | Longest period of depression |
| 4620 | Number of depression episodes |
| 4631 | Ever unenthusiastic/disinterested for a whole week |
| 5375 | Longest period of unenthusiasm / disinterest |
| 5386 | Number of unenthusiastic/disinterested episodes |
| 4642 | Ever manic/hyper for 2 days |
| 4653 | Ever highly irritable/argumentative for 2 days |
| 6156 | Manic/hyper symptoms |
| 5663 | Length of longest manic/irritable episode |
| 5674 | Severity of manic/irritable episodes |
| 6145 | Illness, injury, bereavement, stress in last 2 years |

**Supplemental Table 2.** CVDs and corresponding ICD-10 codes used in this study.

| Data Field 41202 | |
|---|---|
| Diagnoses — main ICD10 | Cardiovascular diseases |
| I48, I481, I482, I483, I484, I489, K621, K622, K623, K624 | Atrial fibrillation/flutter |
| I470, I472, I490, I460, I461, I469, K576, K641, X503, X504, X508, X509 | Ventricular arrhythmias |
| I440, I441, I442, I443, I445, I495, K60, K601, K602, K603, K604, K605, K606, K608, K609, K61, K611, K612, K613, K614, K615, K616, K618, K619 | Bradyarrhythmias |
| I200, I201, I208, I209 | Angina pectoris |
| I210, I211, I212, I213, I214, I219 | Acute myocardial infarction |
| I220, I221, I228, I229 | Subsequent myocardial infarction |
| I231, I232, I233, I236, I238 | Certain current complications following acute myocardial infarction |
| I240, I241, I248, I249 | Other acute ischemic heart diseases |
| I250, I251, I252, I253, I254, I255, I256, I258, I259 | Chronic ischemic heart disease |
| Z951, Z954, Z955, Z958 | Presence of cardiac and vascular implants and grafts |
| I110 | Hypertensive heart disease |
| I130, I132 | Hypertensive heart and renal disease |
| I500, I501, I509 | Heart failure |

**Supplemental Table 3.** Definition of ML performance metrics.

| ML performance metric | Definition |
|---|---|
| True positive (TP) | An instance in which the model correctly predicts the positive class, i.e., the model infers that an individual has established CVD (positive class) for a patient that has established CVD. |
| False positive (FP) | An instance in which the model mistakenly predicts the positive class, i.e., the model infers that an individual has established CVD for a patient has not established CVD. |
| True negative (TN) | An instance in which the model correctly predicts the negative class, i.e., the model infers that an individual has not established CVD for a patient that has not established CVD. |
| False negative (FN) | An instance in which the model mistakenly predicts the negative class, i.e., the model infers that an individual has not established CVD for a patient that has established CVD. |
| Accuracy | The fraction of predictions for which a classification model returns the correct answer, based on the formula: $\text{Accuracy} = \frac{\text{TP+TN}}{\text{TP+TN+FP+FN}}$ |
| Precision | The frequency with which a model was correct when predicting the positive class, i.e., $\text{Precision}= \frac{\text{TP}}{\text{TP+FP}}$ |
| Recall | The proportion of positive identifications that was actually correct, i.e., $\text{Recall}= \frac{\text{TP}}{\text{TP+FN}}$ |

**Supplemental Table 4**. Bone diseases and corresponding ICD-10 codes used in this study.

| **Data Field 41202** | | |
|---|---|---|
| **Top level description** | **Bone Diseases** | **Diagnoses—main ICD10** |
| A15–A19 Tuberculosis | Tuberculosis of bones and joints | A18.0 |
| B90-B94 Sequelae of infectious and parasitic diseases | Sequelae of tuberculosis of bones and joints | B90.2 |
| C40 Malignant neoplasm of bone and articular cartilage of limbs | Scapula and long bones of upper limb | C40.0 |
| | Short bones of upper limb | C40.1 |
| | Long bones of lower limb | C40.3 |
| | Overlapping lesion of bone and articular cartilage of limbs | C40.8 |
| | Bone and articular cartilage of limb, unspecified | C40.9 |
| C41 Malignant neoplasm of bone and articular cartilage of other and unspecified sites | Bones of skull and face | C41.0 |
| | Mandible | C41.1 |
| | Vertebral column | C41.2 |
| | Ribs, sternum, and clavicle | C41.3 |
| | Pelvic bones, sacrum, and coccyx | C41.4 |
| | Bone and articular cartilage, unspecified | C41.9 |
| D16 Benign neoplasm of bone and articular cartilage | Scapula and long bones of upper limb | D16.0 |
| | Short bones of upper limb | D16.1 |
| | Long bones of lower limb | D16.2 |
| | Short bones of lower limb | D16.3 |
| | Bones of skull and face | D16.4 |
| | Lower jaw bone | D16.5 |
| | Vertebral column | D16.6 |
| | Ribs, sternum, and clavicle | D16.7 |
| | Pelvic bones, sacrum, and coccyx | D16.8 |
| | Bone and articular cartilage, unspecified | D16.9 |
| D48 Neoplasm of uncertain or unknown behavior of other and unspecified sites | Bone and articular cartilage | D48.0 |
| M80–M85 Disorders of bone density and structure | Osteoporosis with pathological fracture | M80 |
| | Osteoporosis without pathological fracture | M81 |
| | Osteoporosis in diseases classified elsewhere | M82 |
| | Adult osteomalacia | M83 |
| | Disorders of continuity of bone | M84 |
| | Other disorders of bone density and structure | M85 |
| M86–M90 Other osteopathies | Osteomyelitis | M86 |
| | Osteonecrosis | M87 |
| | Paget's disease of bone [osteitis deformans] | M88 |
| | Other disorders of bone | M89 |
| | Osteopathies in diseases classified elsewhere | M90 |
| M91–M94 Chondropathies | Juvenile osteochondrosis of hip and pelvis | M91 |
| | Other juvenile osteochondrosis | M92 |
| | Other osteochondropathies | M93 |
| | Other disorders of cartilage | M94 |

| | | |
|---|---|---|
| M95–M99 Other disorders of the musculoskeletal system and connective tissue | Other acquired deformities of musculoskeletal system and connective tissue | M95 |
| | Postprocedural musculoskeletal disorders, not elsewhere classified | M96 |
| | Biomechanical lesions, not elsewhere classified | M99 |

**Supplemental Figure 1.** ML model predictions of bone disease from CVD training data.

Accuracy, precision, and recall results from the ensemble machine learning model and all five individual approaches to predict bone disease from the combined traditional CVD risk and psychological factors training data.

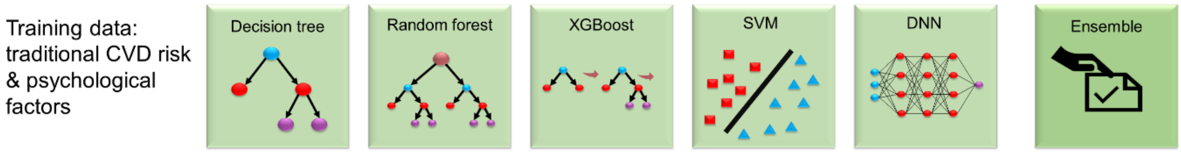


| | Decision tree | Random forest | XGBoost | SVM | DNN | Ensemble |
|---|---|---|---|---|---|---|
| **Accuracy** | **52.59%** | **50.99%** | **51.71%** | **53.52%** | **53.31%** | **54.32%** |
| Precision | 60.06% | 56.58% | 64.42% | 61.33% | 60.54% | 53.28% |
| Recall | 52.86% | 51.70% | 51.92% | 53.64% | 53.50% | 61.32% |

**Supplemental Figure 2.** Date of first CVD in-patient diagnosis versus date of completing UK Biobank mental health questionnaire (started in August 2016).

A) All Biobank data from 1996. B) Expansion of the purple box in (A). highlighting the CVD diagnosis period that overlaps with availability of the mental health questionnaire; only patients to the right of the red dotted line completed the mental health questionnaire before receiving their CVD diagnosis.

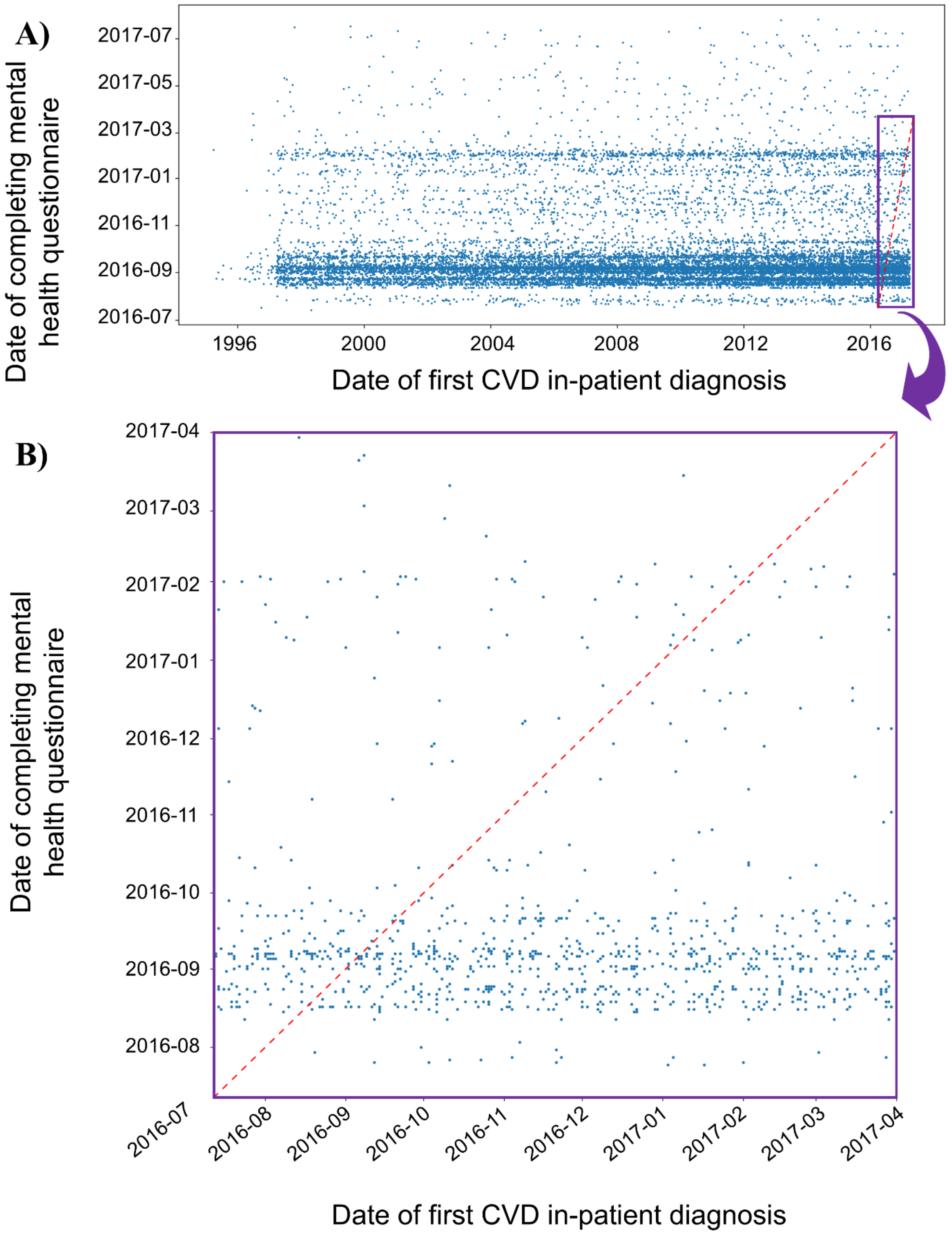
